# Bridging the Gap: Identifying Barriers and Strategies for Widespread Implementation of Long-acting Injectable Antiretroviral ART in Sub-Saharan Africa: A Scoping Review

**DOI:** 10.1101/2024.11.21.24317714

**Authors:** Pierre Gashema, Patrick Gad Iradukunda, Eric Saramba, Tumusime Musafiri, Thérèse Umuhoza, Felix Ndahimana, Angelique Ingabire, Moise Mukire Ndoli, Gerard Mutagoma, Ivan Emil Mwikarago, Eric Nyirimigabo, Muhayimpundu Ribakare, Jean de Dieu Harelimana, Enos Moyo, Tafadzwa Dzinamarira, Claude Mambo Muvunyi

## Abstract

**Background:** Long-acting injectable antiretroviral therapy (LAI ART) presents a new approach to HIV treatment and addressing the challenges of daily oral ART in Sub-Saharan Africa (SSA) remains paramount. This review collates existing literature on barriers and strategies for implementing LAI ART in the SSA region, identifying knowledge gaps and research priorities.

**Methods:** We performed a thorough literature search, encompassing electronic databases and grey literature sources. Our review included 18 studies published between 2014 and 2023, focusing on the acceptability, feasibility, effectiveness, and cost-effectiveness of LAI ART in SSA. Data extraction involved study characteristics, population, intervention, outcomes, and context, with findings synthesized using a narrative approach.

**Results:** The results indicate high demand and acceptability of LAI ART among people living with HIV in SSA, particularly those facing stigma and discrimination. LAI ART can improve adherence, retention, and viral suppression while reducing pill burden and frequent clinic visits. Implementation challenges include lack of regulatory approval, high cost, limited supply chain, health system capacity, trained staff, and cold storage facilities. Further evidence on safety and efficacy, advocacy, policy, and community engagement are needed to ensure accessibility and equity.

**Conclusion:** Knowledge gaps and research priorities include dosing regimens, long-term outcomes, drug resistance, impact on sexual and reproductive health, interactions with other medications and co-infections, and preferences and experiences of different subgroups.

## 1. INTRODUCTION

Over four decades have passed since the initial documented case of HIV[1]. In 2022, approximately 39 million individuals were reported to be living with HIV, and 1.3 million new cases of HIV/AIDS were recorded [2]. The specter of HIV/AIDS continues to loom large in Sub-Saharan Africa (SSA), disproportionately impacting millions of lives. Since its introduction in the late 1980s, antiretroviral therapy (ART) has revolutionized the field of HIV treatment [3]. Current ART regimens are extremely effective, well-endured, and safe, frequently consisting of a single multi-drug tablet taken once daily [4]. According to the 2022 global HIV statistics, approximately 29.8 million individuals were receiving ART [2].

Antiretroviral therapy (ART) has undeniably transformed the landscape, yet substantial gaps remain in achieving optimal viral suppression and treatment goals [5]. One transformative contender on the horizon is long-acting injectable (LAI) ART, which holds the potential to revolutionize HIV care by minimizing the burden of daily medication adherence [3]. Several injectable agents are in different stages of development, including cabotegravir, rilpivirine, lenacapavir, islatravir, albuvirtide, and ibalizumab [1] In 2021, Cabotegravir/Rilpivine (CAB/RPV), the initial long-acting injectable antiretroviral medication (LAI ART), was licensed for sustained HIV-1 treatment for people living with HIV (PLWH), [6]. For PLWH, a LAI ART is considered a substitute for the daily oral regimen. Current daily oral ART, though effective, faces hurdles – from stigma and forgetfulness to complex dosing schedules and drug-drug interactions. These challenges contribute to suboptimal adherence, jeopardizing viral suppression and ultimately, the health and well-being of PLWH [5]. LAI ART, with its extended dosing intervals and simplified administration, offers a promising solution [3]. This review meticulously dissects the current knowledge base on LAI ART in SSA, drawing insights from diverse sources: research studies, policy documents, and stakeholder perspectives.

A study carried out in the United States and Spain among PLWH revealed a preference for LAI ART regimens over daily oral ART [7]. In the same study, healthcare professionals expressed concerns about resistance and the complex clinical supervision associated with LAI ART medication over oral ART. This risk stems from the prolonged existence of LAI ART in the patient’s body, potentially posing challenges if the prescription is discontinued [7]. Furthermore, in a clinical trial examining the use of LAI ART as a substitute for daily oral ART, the study findings demonstrated that individuals in the LAI ART groups were significantly higher satisfied with therapy in comparison to patients receiving oral ART [4].

This scoping review aimed to assess the landscape surrounding LAI ART in SSA, delving into the challenges and opportunities that pave the road to its potential large-scale implementation. By illuminating these challenges and opportunities, this review aims to equip policymakers, healthcare providers, researchers, and advocates with a comprehensive understanding of the LAI ART landscape in SSA. Ultimately, we strive to pave the way for informed decision-making and strategic interventions that can unlock the full potential of LAI ART and bring us closer to a future where HIV is truly controlled and managed, one injection at a time. This review, therefore, stands as a critical step towards harnessing the transformative power of LAI ART and propelling SSA toward a future unshackled by the chains of HIV/AIDS.

## 2. METHODOLOGY

### a. Study design, Information sources and literature search

This scoping review adapted the guidelines for the systematic scoping review. The sources of information were PubMed, Google Scholar, Web of science and EMBASE databases. The main search terms included “People Living with HIV”, “Sub-Saharan Africa”, “Long-Acting Injectable”, “Antiretroviral” and “Challenges and Opportunities”. All database searches were for sources from 2014 to 2023. To maximize the inclusion criteria, the reference lists of all full-text screening articles were searched for relevant studies. To ensure the consistency and quality of the findings, two independent authors searched for the articles and the research team screened and synchronized the extracted data.

### b.Research questions

This scoping review sought to answer the following questions:

i. How acceptable and feasible is LAI ART implementation in SSA?
ii. What are the challenges of LAI ART implementation in SSA?
iii. What are the possible solutions to LAI ART implementation challenges in SSA?
iv. What are the potential benefits of LAI ART implementation in SSA?

### c. Study selection and inclusion criteria

In this review, we included studies conducted in SSA from 2014 to 2023 and reported the use of LAI ART and the associated challenges. The studies selected focused on the challenges and opportunities of scaling up LAI ART in SSA, provided an overview of the current state of LAI ART implementation in SSA, barriers, and facilitators to its expansion, and discussed the outcomes/results for clinical trials conducted on LAI ART. Studies that included primary data on perceived challenges and acceptability, feasibility, and cost-effectiveness of LAI ART, commentary, review, and perspectives papers were included. Papers reporting the same findings which are outside of Africa were not included in the review.

### d. Data abstraction and screening process

Our screening procedure was guided by Arksey and O’Malley’s framework [8] in all three stages: Title, abstract, and full-text. Three reviewers conducted independent title screening which allowed the research team to come up with an overage of studies before starting the abstract, full-text screening and hand-searching were performed in all articles that were eligible for full-text screening. We conducted data abstraction after the identification of the final studies to be included in the review. Data abstraction form contained article titles, authors, year of publication, countries, target populations, study design, key findings on the use of LAI ART, accessibility and usability of LAI ART, perspectives of end users on LAI ART, cost-effectiveness of LAI ART, and recommendation to program people when LAI ART is going to be rolled out in the African setting (All this information were removed in the form to allow blind peer review).

### e. Data synthesis

Both quantitative and non-quantitative outcomes were collected and tabulated for each study. Thereafter, collating, summarizing, and reporting the findings were done. The findings of the study were reported in a narrative synthesis.

## 3. RESULTS

At the initial stage of searching 513 articles were retrieved. 50 articles were eligible for abstract screening. A total of 30 articles were included after abstract screening, and 12 articles were excluded at this stage. 30 articles underwent full-text screening (supplementary file 1 of articles eligible for full-text screening). Twelve studies were excluded at this stage since they were conducted outside of Africa. A total of 18 articles were included in the final review. However, the study by Castor et al. (2020) was included in the review because study areas were not specified, and studies by Mantsios et al. (2022) [9] and Deanna Kerrigan et al. (2020) [10] were included in the final review because the studies were multi-country. Screening details are presented in Figure 1 below, which is normally used to report items for systematic review (PRISMA).

**Figure 1.**
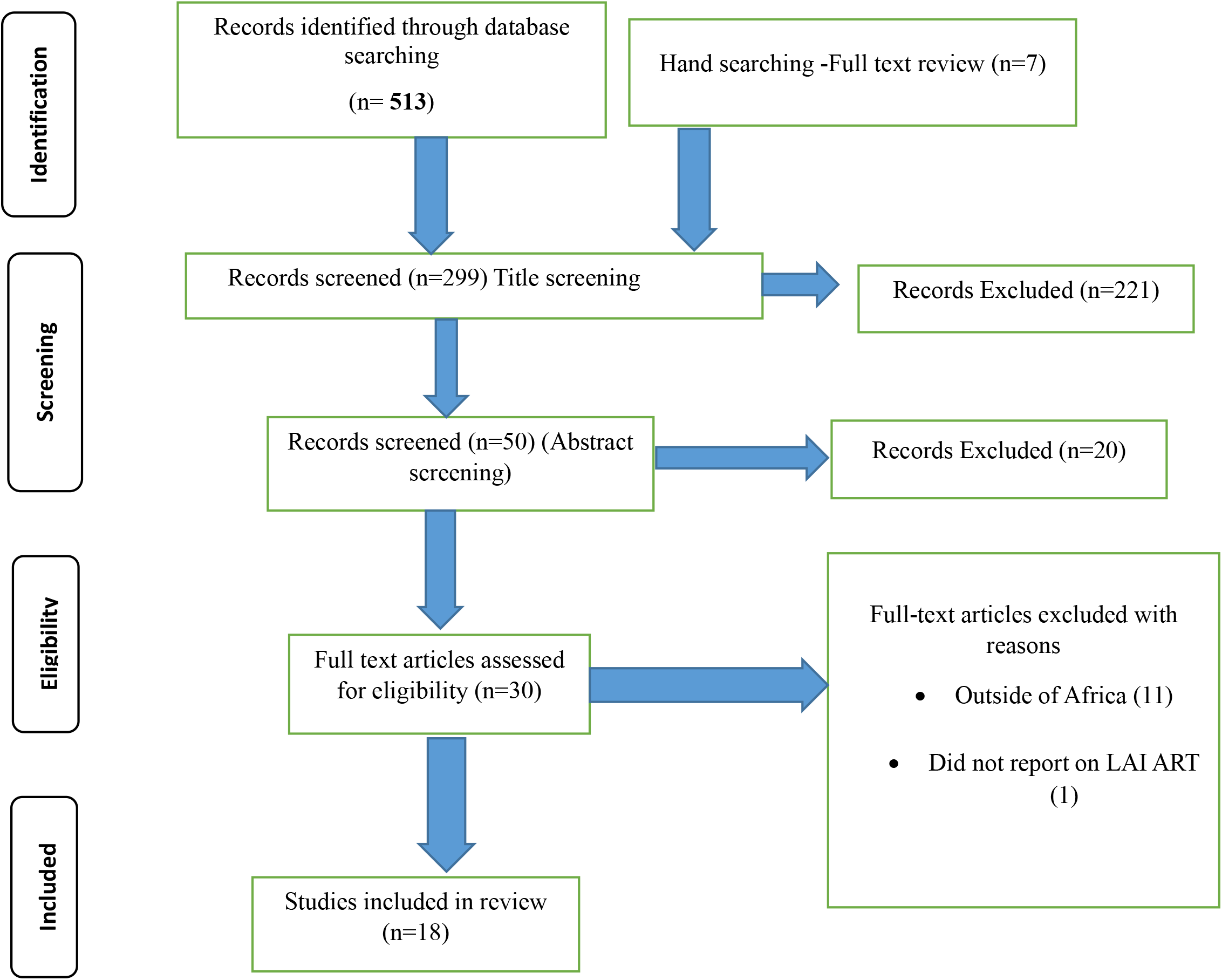
Details of screening procedure.

### a) Characteristics of included studies

The literature search yielded a variety of studies: four qualitative studies, one quantitative study, five review studies, three cross sectional studies, two mixed-methods studies, one modelled economic evaluation and threshold analysis, one commentary, and one mathematical modeling study. The studies covered different countries and populations in SSA including Tanzania, South Africa, Uganda, Kenya and Guinea-Bissau. This review encompasses a diverse array of populations affected by or at risk of HIV infection in various regions. Included groups are female sex workers living with HIV in Tanzania and the Dominican Republic [10], HIV-negative sexually active heterosexual men in Johannesburg [11], infants, children, and adolescents at risk of HIV [12], as well as a broad spectrum of individuals such as adolescents, young adults [13], and key populations in different contexts. The study also considers pregnant and postpartum women in oral PrEP studies, individuals with virological failure on first-line antiretroviral therapy, and a general category of people living with HIV [1]. and had different intervention and outcome characteristics. The studies also had diverse and sometimes conflicting findings on the challenges and opportunities of LAI ART implementation in SSA.

### b) Study findings

#### i. Acceptability and feasibility of LAI ART

The acceptability of LAI ART in SSA is influenced by various factors at the individual, population, and health system levels. Several studies [11] have explored the demand, preferences, and experiences of different groups of PLWH in SSA regarding LAI ART. These studies have shown that LAI ART has a high potential acceptability among PLWH, especially among those who face stigma, discrimination, or violence due to their HIV status or other factors. For example, a study in South Africa found that LAI PrEP was highly acceptable among heterosexual men in urban settings, who valued its convenience and discretion [11]. Another study in Kenya reported positive initial reactions to LAI ART among women and adolescents, who appreciated its potential to improve their health outcomes and quality of life [14]. A study in Tanzania found high demand and acceptability for LAI ART among female sex workers (FSW), influenced by various factors such as quality of HIV care, clinic access, income, and years on ART [10]. However, another study in South Africa found low preference for LAI ART over oral ART among a general sample of PLWH, influenced by factors such as medication stock-outs, side-effects, pill-burden, treatment changes, and HIV stigma [15].

#### ii. Challenges revealed from included studies

Several studies in Africa, explored the use of LAI ART for HIV prevention and treatment, and found that it had the potential to improve the lives of people living with or at risk of HIV. However, they also faced various challenges that could limit their effectiveness and accessibility. One study in South Africa reported that LAI ART was more costly than oral ART, and could cause drug resistance and side effects [13]. A review study revealed potential challenges LAI ART might face during implementation, such as the lack of regulatory approval, the high cost, the limited supply chain, and the need for cold storage and trained staff [1]. A study in SSA reported that long-acting extended-duration (LAED) formulations, such as implants, injectables, and vaginal rings, faced the challenges of shortage of healthcare providers, inadequate infrastructure, patient concerns, cost, and drug resistance [16]. A fourth study among PLWH in low- and middle-income countries (LMICs) reported that long-acting HIV treatment and prevention such as injectable antiretrovirals and broadly neutralizing antibodies, faced the challenges of acceptability, demand, supply, delivery, monitoring, and cost-effectiveness. Other study identified the health system and client challenges that CAB-LA faces in Africa, such as cost, refrigeration, training, knowledge, accessibility, side effects, stigma, and support [17] In Kenya, potential side effects were a particular concern among women and most participants preferred clinic-based administration over self-injections at home due to concerns about safety, privacy, and potential need for refrigeration [14]. These studies showed that LAI ART had the potential to transform the lives of millions of people affected by HIV in Africa, but also that there were many challenges and opportunities that needed to be considered and addressed.

#### iii. Proposed solution to identified challenges

In Tanzania and the Dominican Republic, a study suggested that FSW, who are among the most vulnerable groups to HIV, could benefit from LAI ART if they receive community-driven approaches that include tailored health education, improved patient-provider communication and quality of care, and strategies to facilitate appointment adherence [10]. These approaches could help FSW overcome the barriers of stigma, discrimination, violence, and lack of access that often prevent them from using oral ART.

The study also estimated that the cost per CAB-LA injection would need to be between $9.03 and $14.47 for it to be similarly or more cost-effective than daily oral tenofovir disoproxil fumarate and emtricitabine (TDF/FTC), and hence acceptable to the South African government [13].

In SSA, a study evaluated the feasibility and acceptability of long-acting HIV treatment and prevention (LAHTP), such as injectable antiretrovirals and broadly neutralizing antibodies, and highlighted the need for decision-makers to define and gather relevant data to inform their investment case within the existing health systems context. The study also emphasized the importance of engaging with key stakeholders, including PLWH, providers, regulators, and funders, to ensure that LAHTP meets the needs and preferences of the end-users and is aligned with the national and global HIV goals.

In Uganda, a study examined the factors influencing ART adherence among youth living with HIV, and revealed that programs and policies to improve ART adherence should address the specific challenges faced by this age group, such as economic independence, school support, family and peer engagement, and medication simplification [18]. These factors could affect the willingness and ability of youth to use LAI ART, which could offer them more convenience and privacy than oral ART.

In SSA, one study proposed that the region should develop laboratory capabilities, enhance research, train and retain more healthcare providers, invest in infrastructure, integrate services, advocate for patent waivers, and procure drugs collectively, in order to improve the availability and affordability of LA-ART. The study also suggested that advocating for waiving of CAB-LA patent licence, conducting demonstration projects in Africa, promoting the use of renewable energy sources, healthcare provider training, task shifting, community engagement, client education, and implementing adherence promotion strategies, could facilitate the uptake and effectiveness of LA ART [16]. Another study highlighted the differences between HIV treatment and prevention programs, and the need to consider the building blocks of differentiated service delivery (DSD): who (provider), where (location), when (frequency) and what (package of services). The study also suggested that LAED regimens should leverage DSD models that emphasize access at the community level and self-management, and that address the specific barriers and needs of the affected populations [19].

#### iv. Potential Benefits associated with LAI-ART

A global study highlighted the potential benefits of long-acting products for improving adherence, reducing stigma, and enhancing quality of life for infants, children, and adolescents affected by HIV [12]. Another study on PLWH and at-risk individuals for HIV infection reviewed the advantages of injectable antiretrovirals, such as longer duration of action, more stable drug levels, less frequent administration, and improved adherence and retention. A study in Africa evaluated the feasibility and acceptability of long-acting injectable cabotegravir (CAB-LA) for HIV-1 prevention, and found that it was a promising option for reducing the risk of HIV transmission [20]. A study in Kenya estimated the cost-effectiveness and impact of LAI ART on HIV incidence and mortality among adolescents and young adults over 10 years, compared to oral ART. The study found that LAI ART could prevent more new HIV infections and deaths than oral ART, and that the cost threshold for LAI ART to be cost-effective was lower when non-adherent oral ART users were assumed to be less likely to switch to LAI ART [21]. A study in South Africa compared the benefits and issues of long-acting extended-duration (LAED) regimens for HIV treatment and prevention and suggested that LAED regimens offer unique benefits for expanding uptake, effective use and adherence.

## 4 DISCUSSION

Our review highlighted that LAI ART can also improve adherence, retention, and viral suppression compared to oral ART, and reduce the pill burden and the frequency of clinic visits. Adherence to HIV treatment was the main topic over the last three decades and studies reported adherence rate ranging from 50% to 90% depending on the measurement methods and the settings [22], Which is in line with our findings linked with the convenience and preference of LAI ART over oral ART, the reduced stigma and discrimination associated with LAI ART, and the improved quality of life and satisfaction with LAI ART[23], [24]. Report primary data and study population also led to the differences in the findings, as some studies focused on specific subgroups of PLWH, such as men who have sex with men, transgender women, or people who inject drugs, who may face different barriers and facilitators to adherence [25]–[28].

This review revealed potential challenges LAI ART might face during implementation, such as the lack of regulatory approval, the high cost, the limited supply chain, and the need for cold storage and trained staff. This was also reported in the study conducted by Jolayemi et al. (2022) who explored the perspectives of consumers, clinical and non-clinical stakeholders on LAI ART in Los Angeles County, California [29]. They found that regulatory approval remains an issue before rolling out this lifetime treatment, as the first LAI ART product was approved by the U.S. Food and Drug Administration (USFDA) in January 2021 and others are in the treatment (and prevention) pipeline [29]. Cost effectiveness and challenges in supplying LAI ART was reported by other scholars, Reza et al. (2013), who developed a three-step approach to evaluate and prioritize the cost-effectiveness criteria in supply chain management using fuzzy multiple attribute decision-making [30]. They suggested that LAI ART could reduce the total cost of HIV treatment by improving adherence and reducing the frequency of clinic visits, but also highlighted the need for adequate information, education, and counseling, as well as supportive relationships and follow-up visits [31].

This review highlighted that there is inadequate health system capacity, the need for trained staff and cold storage facilities, the uncertainty about the safety and efficacy of LAI ART in different populations and settings, and the potential for drug resistance and adverse events [25], [32]. These findings were also reported in the study done by Margolis et al. (2015) [33], who summarized the current state and future directions of LAI ART research and development. Studies noted that infrastructure was the main issue and a need for capacity building for staff were also common challenges associated with LAI ART distribution among the general population [34]. Another study also emphasized the importance of addressing the gaps in knowledge and evidence on the long-term outcomes and safety of LAI ART, especially in resource-limited settings and diverse populations [35]

The review suggested that to overcome these barriers, there is a need for more evidence on the safety and efficacy of LAI ART in SSA, as well as for more advocacy, policy, and community engagement to ensure the accessibility and equity of LAI ART. The suggested intervention to overcome barriers was also reported in the pilot study done by Christopoulos et al. (2023) in the USA, who demonstrated promising early treatment outcomes with LAI ART among people who use drugs, a population that faces greater challenges to adherence and retention [34]. Community engagement is always discussed in HIV programs, and the theme for World AIDS Day 2023 is “Let Communities Lead” [36]. It has been seen that community engagement is key for HIV intervention and LAI ART will be successfully distributed to the end users via community mobilization and empowerment [37].

The review highlighted the knowledge gaps and research priorities for LAI ART in SSA, such as the optimal dosing regimen, the long-term outcomes, the drug resistance, the impact on sexual and reproductive health, the interaction with other medications and co-infections, and the preferences and experiences of different subgroups of PLWH. These concerns were also reported by Lahuerta et al. (2013) [38]. In their review of the current state and future directions of LAI ART research and development, Kanazawa et al. (2021) noted that the optimal dosing regimen of LAI ART may vary depending on the pharmacokinetics, pharmacodynamics, and pharmacogenetics of different populations and settings [23]. Drug resistance is a major challenge for the sustainability and effectiveness of LAI ART, as the long half-life of the drugs may increase the risk of selecting resistant strains in cases of suboptimal adherence or treatment interruption [39].

The impact of LAI ART on sexual and reproductive health is another important area of research, as LAI ART may affect the hormonal contraception, pregnancy, breastfeeding, and sexually transmitted infections among PLWH [3]. The interaction of LAI ART with other medications and co-infections is also a critical issue, as many PLWH may have comorbidities or require concurrent treatments that may alter the pharmacokinetics or pharmacodynamics of LAI ART [35]. Finally, the preferences and experiences of different subgroups of PLWH are essential to understand the acceptability, feasibility, and satisfaction of LAI ART, as well as the potential barriers and facilitators to its uptake and adherence. These findings were in agreement with other studies that explored the perceptions and attitudes of various stakeholders towards LAI ART [39]. Long-term outcomes and drug resistance are the main issues among PLWH and need special attention for HIV control programs [40].

## 5 Limitations

This review has some limitations and strengths. Even though the search retrieved different studies addressing the issue of the availability of LAI ART, one of the limitations is linked to our electronic search which might have missed out some articles. Another limitation of this review is associated with the articles included, which were only in English, and there is potential of missing non-English articles. And also, the review did not follow the steps for a systematic review and this may increase bias when deciding which article to be included or not. However, we believe that this review presents important information to guide program people and what can be done before rolling out LAI ART in SSA.

## 6 Conclusion

This review focuses on the feasibility and acceptability of LAI ART in SSA, where high HIV prevalence and oral ART adherence challenges exist. The objectives include identifying influencing factors, analyzing implementation challenges and opportunities, proposing solutions, and evaluating potential benefits for health outcomes. The review findings suggest a potential high acceptability, especially among those facing stigma. LAI ART could enhance adherence, but challenges like cost and regulatory approval need consideration. Despite limitations, the review provides a comprehensive overview, highlights gaps, proposes solutions, and underscores LAI ART’s potential benefits in preventing new infections, emphasizing the ongoing effort to end the HIV epidemic by 2030.

## Data Availability

All the data we used is available in the supplementary files.

## 7 List of Abbreviation

AIDS: Acquired Immune Deficiency Syndrome
ART: Antiretroviral therapy
CAB/RPV: Cabotegravir/Rilpivine
DSD: Differentiated Service Delivery
FSW: Female Sex Workers
HIV: Human Immunodeficiency Virus
LAED: Long-acting extended-duration
LAHTP: Long-acting
HIV: treatment and prevention
LAI ART: Long-acting injectable antiretroviral therapy
LMICs: low- and middle-income countries
PLWH: People Living with HIV
PrEP: Pre-exposure prophylaxis
SSA: Sub-Saharan Africa
USFDA: U.S. Food and Drug Administration

## Declaration

## Ethical Consideration

Not Applicable

## Consent for publication

Not Applicable

## Clinical Trials Number

Not Applicable

## Availability of data and materials

All the data we used is available in the supplementary files.

## Authors’ contributions

PG contributed to the design of the work, analysis, interpretation of results, drafting, and revision of the manuscript. PGI, ES, MMK, FN,AI,GM, MR, ER, TM and TU contributed to the design of the work and Screening of the papers. IEM, JDH, EM, TD and CMM contributed to the design of the work, interpretation of results, and revision of the manuscript.

## Acknowledgement

All authors acknowledged contribution of this work

---

**Supplementary file 1: List of paper included in Final Review**

**Supplementary file 2: Table Summarizing Keys findings**

**Supplementary File 3: PRISMA Check list**

