## Supplemental file 1 for "Bridging the Gap: Identifying Barriers and Strategies for Widespread Implementation of Long-acting Injectable Antiretroviral ART in Sub-Saharan Africa: A Scoping Review"

### SUPPLEMENTARY FILE 2: KEY FINDING FROM PAPER RETAINED FOR FINAL REVIEW

| Author & Year | Title | Findings | Recommendations | DOI |
|---|---|---|---|---|
| Deanna Kerrigan et al.1(2020) | "A dream come true": Perspectives on long-acting injectable antiretroviral therapy among female sex workers living with HIV from the Dominican Republic and Tanzania | Most FSW from the DR and Tanzania were very likely to use LA ART if available. In Tanzania, better HIV care and communication increased LA ART likelihood. In the DR, easier clinic access and more income from sex work did. In both, longer ART use also did. Qualitative findings showed LA ART could address sex work barriers to daily oral ART. | The study suggested community-driven approaches which include tailored health education and improved patient-provider communication and quality of care, as well as strategies to facilitate appointment adherence are needed to optimize LA ART use among FSW. | 10.1371/journal.pone.0234666 |
| Chih-Yuan Cheng et al. (2019) | Determinants of heterosexual men's demand for long-acting injectable pre-exposure prophylaxis (PrEP) for HIV in urban South Africa | Almost half of participants preferred LAI PrEP, a third oral PrEP, and a fifth condoms. Having children, higher risk attitude, and longer ART use increased LAI PrEP preference. Having unprotected anal sex and valuing STI over HIV prevention decreased it. | The study found that LAI PrEP was highly demanded and accepted by heterosexual men in urban South Africa. It urged HIV prevention to account for their diverse preferences and traits. It also suggested community-based methods to enhance health education, communication and care for LAI PrEP users. | 10.1186/s12889-019-7276-1 |

**SUPPLEMENTARY FILE 2: KEY FINDING FROM PAPER RETAINED FOR FINAL REVIEW**

| | | | | |
|---|---|---|---|---|
| Elaine J Abrams et al. (2020) | Potential of Long-Acting Products to Transform the Treatment and Prevention of Human Immunodeficiency Virus (HIV) in Infants, Children, and Adolescents | The article highlighted the potential benefits of long-acting products for improving adherence, reducing stigma, and enhancing quality of life for infants, children, and adolescents affected by HIV. The article also discussed the pharmacokinetic, safety, and ethical issues that need to be addressed before these products can be widely implemented in pediatric HIV care. | The article argued for long-acting products for HIV prevention and treatment in children. It called for collaboration among stakeholders to develop and deliver these products. It also advised to combine long-acting products with HIV services that suit children's needs and preferences | 10.1093/cid/ciac754 |
| Jamieson et al., 2022 | Relative cost-effectiveness of long-acting injectable cabotegravir versus oral pre-exposure prophylaxis in South Africa based on the HPTN 083 and HPTN 084 trials: a modelled economic evaluation and threshold analysis | Long-acting injectable cabotegravir (CAB-LA) prevented more HIV infections than oral PrEP but cost more. The cost per infection prevented was \$6053–6610 (oral PrEP) and \$4471–6785 (CAB-LA). | The cost per long-acting injectable cabotegravir injection would need to be between \$9·03 and \$14·47 for it to be similarly or more cost-effective than daily oral tenofovir disoproxil fumarate and emtricitabine, and hence acceptable to the South African Government. | 10.1016/S2352-3018(22)00251-X |
| Jespersen et al., 2020 | HIV treatment in Guinea-Bissau: room for improvement and time for new treatment options | The HIV cohorts in Guinea-Bissau are unique research platforms and reflect many African countries. They face many challenges in HIV testing and treatment, making the "90–90–90″ goal1 unachievable by 2020. Keeping viral loads undetectable for a cure seems impossible now. | Long-acting antiretroviral treatment options like injections or implants may suit Guinea-Bissau better and help cure HIV. Whether sub-Saharan Africa should improve HIV treatment by using existing or new options is unclear. | 10.1186/s12981-020-0259-6 |

### SUPPLEMENTARY FILE 2: KEY FINDING FROM PAPER RETAINED FOR FINAL REVIEW

| | | | | |
|---|---|---|---|---|
| Castor et al.,2020 | The only way is up: priorities for implementing long-acting antiretrovirals for HIV prevention and treatment | LAHTP can address some of the limitations of daily oral therapy and improve adherence, retention, and outcomes for PLHIV in LMICs. However, LAHTP also poses individual-level, population-level, and health systems-level challenges, such as acceptability, demand, supply, delivery, monitoring, and cost-effectiveness. | To implement and improve LAHTP, decision-makers need relevant data for their investment case within the health systems context. They also need to involve key stakeholders, such as PLHIV, providers, regulators, and funders, to make sure LAHTP suits the end-users and matches the HIV goals. | 10.1097/coh.0000000000000601 |
| MacCarthy et al. (2018) | How am I going to live?": exploring barriers to ART adherence among adolescents and young adults living with HIV in Uganda | Four barriers to ART adherence emerged: poverty, school attendance, family support, and medication burden. Peer influence was a positive factor for adherence. Disclosure issues were common across the barriers. | Programs and policies to improve ART adherence among youth in Uganda should address the specific challenges faced by this age group, such as economic independence, school support, family and peer engagement, and medication simplification. | 10.1186/s12889-018-6048-7 |
| Toska et al. (2023) | Factors Associated with Preferences for Long-Acting Injectable Antiretroviral Therapy Among Adolescents and Young People Living with HIV in South Africa | 12% of participants preferred long-acting injectable antiretroviral therapy (LAART) over pill regimens. Six factors were associated with LAART preference: medication stock-outs, experiencing side-effects, pill-burden, past-year treatment changes, any HIV stigma, and recent ART initiation. | Adding LAART to existing treatment options for adolescents and young people living with HIV, particularly higher risk groups, would support them to attain and sustain viral suppression and reduce their risk of AIDS-related mortality. | 10.1007/s10461-022-03949-2 |
| Simoni et al. (2021) | "Lighten This Burden of Ours": Acceptability and Preferences Regarding Injectable Antiretroviral Treatment Among Adults and Youth Living With HIV in Coastal Kenya | Many liked the idea of LAI-ART for better adherence, less pills, and less stigma. Women worried about side effects. Most wanted clinic injections over home injections for safety, privacy, and storage. | LAI-ART may be acceptable in Kenya, provided injections are infrequent and delivered in a clinic setting. However, HIV stigma, fear of potential side effects, and limited clinical capacity would need to be addressed. | 10.1177/23259582211000517 |

#### SUPPLEMENTARY FILE 2: KEY FINDING FROM PAPER RETAINED FOR FINAL REVIEW

| | | | | |
|---|---|---|---|---|
| Mgodi et al. (2023) | Advancing the use of Long-Acting Extended Delivery formulations for HIV prevention in sub-Saharan Africa: challenges, opportunities, and recommendations | LAED formulations can ease user burden and adherence issues with oral PrEP and lower HIV risk for vulnerable groups. But they face barriers like lack of providers, infrastructure, patient trust, affordability, and resistance. | Sub-Saharan Africa should develop laboratory capabilities, enhance research, train and retain more healthcare providers, invest in infrastructure, integrate services, advocate for patent waivers, and procure drugs collectively. | 10.1002/jia2.26115 |
| Moyo et al. (2022) | Long-Acting Injectable Drugs for HIV-1 Pre-Exposure Prophylaxis: Considerations for Africa | CAB-LA is a potential HIV-1 prevention option, but has many challenges in Africa, like cost, storage, education, access, effects, stigma, and support. | Some steps to improve CAB-LA access in Africa are: waiving patent licence, doing demonstration projects, using renewable energy, training providers, shifting tasks, engaging community, educating clients, and promoting adherence. | 10.3390/tropicalmed7080154 |
| Wara et al. (2022) | Preferences and Acceptability for Long-Acting PrEP Agents Among Pregnant and Postpartum Women with Experience Using Daily Oral PrEP in South Africa and Kenya | Most preferred injectable PrEP over oral (75%) or ring (87%) for longer effect and privacy. Oral PrEP had side effects and pills. PrEP preferences were long-acting, effective, safe, and free. | Oral PrEP-experienced pregnant and postpartum women expressed a theoretical preference for long-acting injectable PrEP over other modalities, demonstrating potential acceptability among a key population who must be at the forefront of injectable PrEP rollout. | 10.1101/2022.10.29.22281701 |

### SUPPLEMENTARY FILE 2: KEY FINDING FROM PAPER RETAINED FOR FINAL REVIEW

| | | | | |
|---|---|---|---|---|
| Berruti et al. (2020) | Injectable Antiretroviral Drugs: Back to the Future | Injectable antiretrovirals offer benefits over oral drugs, like longer effect, stable levels, less doses, and better adherence. But they have challenges, like cost, storage, delivery, monitoring, safety, resistance, and acceptability. Some injectable drugs are being developed, such as cabotegravir, rilpivirine, lenacapavir, islatravir, albuvirtide, and ibalizumab. | Injectable antiretrovirals could represent a paradigm shift in the management of HIV infection, but they require further research and implementation strategies to overcome the barriers and optimize their use in different settings and populations. | 10.3390/v13020228 |
| ulhane J, et al., 2020 | Modeling the health impact and cost threshold of long-acting ART for adolescents and young adults in Kenya | The findings showed that LA-ART could prevent more HIV infections and deaths than oral ART in 10 years. The annual per-person cost of LA-ART was $89-$236 to be cost-effective under the thresholds of $500-$1,508 per DALY1 averted. The cost threshold was lower with less switching from oral ART. | The authors suggested LA-ART as a possible option for better HIV outcomes in Kenyan youth, especially those with oral ART adherence issues. They also called for more research on LA-ART's safety, efficacy, acceptability, cost-effectiveness, feasibility, and scalability. | 10.1016/j.eclinm.2020.100454 |
| Reynolds Z, McCluskey SM, Moosa MY, et al. (2020) | Who's slipping through the cracks? A comprehensive individual, clinical and health system characterization of people with virological failure on first-line HIV treatment in Uganda and South Africa | More Ugandans (16.3%) than South Africans (10.4%) failed first-line ART. Most (82.9%) were unaware and few (18.4%) switched to second-line ART. Younger age, lower education, unemployment, alcohol, depression, poor adherence, and low clinic attendance increased failure risk. The authors also reported HIV care gaps and | The authors recommended that interventions to improve the detection and management of virological failure should be prioritized and tailored to the specific needs and contexts of the affected populations. They also recommended that more resources and support should be allocated to strengthen the HIV service delivery and quality at the clinic level. | 10.1002/jia2.25587 |

| | | | | |
|---|---|---|---|---|
| | | challenges, like late testing, poor counseling, and limited second-line ART. | | |
| Grimsrud A, et al. (2023) | The importance of the “how”: the case for differentiated service delivery of long-acting and extended delivery regimens for HIV prevention and treatment | The authors said LAED regimens have benefits for HIV uptake, use and adherence, but also new challenges for service delivery and client choices. They showed the differences between HIV treatment and prevention, and the importance of DSD building blocks: who, where, when and what. They advised LAED regimens to use DSD models that stress community access, self-management, and population needs and barriers. | The authors recommended that service delivery and client considerations should be integrated into the development, trial and early implementation of LAED regimens for HIV prevention and treatment, and that more research and advocacy are needed to ensure that these innovative products reach those who most stand to benefit. | 10.1002/jia2.26095 |

### SUPPLEMENTARY FILE 2: KEY FINDING FROM PAPER RETAINED FOR FINAL REVIEW

| | | | | |
|---|---|---|---|---|
| Kim (2021) | Long-Acting Injectable Antiretroviral Agents for HIV Treatment and Prevention | T The author reported that LAI-ARV agents, like cabotegravir and rilpivirine, performed well in trials for HIV treatment and prevention, with high suppression and efficacy, and good safety and tolerability. LAI-ARV agents have benefits over oral drugs, like better adherence, convenience, privacy, and integration. But they have challenges, like regular injections, monitoring, and management of events, resistance, and discontinuation. | The author recommended that LAI-ARV agents should be offered as an additional option for HIV treatment and prevention, as part of a comprehensive package of services that includes HIV testing, counseling, and linkage to care. The author also suggested that further research is needed to address the operational, ethical, and regulatory issues related to LAI-ARV implementation, as well as the preferences and needs of different communities. | 10.1007/s11904-021-00572-5 |
| Mantsios et al. (2022) | "She is the one who knows": A qualitative exploration of oral and injectable PrEP as part of a community empowerment approach to HIV prevention among female sex workers in the Dominican Republic and Tanzania | The authors reported that FSW in both settings wanted PrEP for their work. Most liked injectable PrEP over oral PrEP for less stigma and pill issues. But some liked oral PrEP for no needles, doubts, or pill trust. The authors also learned that FSW valued peer education for PrEP information and support, and stressed FSW choice and autonomy for PrEP option. | The authors recommended that PrEP should be offered as an additional HIV prevention option for FSW, as part of a comprehensive package of services that includes HIV testing, counseling, and linkage to care. They also suggested that further research is needed to address the operational, ethical, and regulatory issues related to PrEP implementation, as well as the preferences and needs of different communities of FSW. | 10.1371/journal.pgph.0000981 |

### SUPPLEMENTARY FILE 2: KEY FINDING FROM PAPER RETAINED FOR FINAL REVIEW

#### Common findings, recommendations, and challenges/gaps among the included studies

The studies explored the potential of long-acting injectable antiretroviral therapy (LAI-ART) or pre-exposure prophylaxis (LAI-PrEP) for HIV prevention and treatment in African settings. They also identified several challenges and gaps, such as cost, storage, delivery, monitoring, safety, resistance, acceptability, and client preferences. They found that LAI-ART or LAI-PrEP could improve HIV outcomes by reducing infections, deaths, stigma, and pill burden, and increasing convenience, privacy, and effectiveness. They recommended further research, community engagement, health education, and service delivery to optimize the development and implementation of LAI-ART or LAI-PrEP.
